# Consumer Opinions, Lot-to-Lot Variability, and Pharmacokinetics of Transdermal Melatonin Products: A Randomized, Crossover Clinical Trial

**DOI:** 10.64898/2026.05.27.26354234

**Authors:** Kathy Bonilla, Vanessa M. Sherman, Alba S. Arbaiza, Mason Dougherty, Lisa E. Olson

**Affiliations:** University of Redlands, Department of Biology, Redlands, CA, USA

**Keywords:** melatonin, transdermal, sleep, pharmacokinetics, saliva, HPLC, consumer opinion

## Abstract

In some countries, melatonin is sold without a physician’s prescription and dosage is unregulated. Transdermal products have become popular including those marketed for children. We measured consumer assumptions about these products among adult residents of the United States, analyzed lot-to-lot variability, and compared the pharmacokinetics of melatonin administered in oral, lotion, and bath product forms. Survey respondents (*n*=199) believed oral melatonin was more effective than transdermal products and that all melatonin products were relatively safe. Melatonin lotion products analyzed by HPLC displayed lot-to-lot variability as well as changes in formulation and product claims. To determine pharmacokinetics, three different treatments (oral tablets, lotion, and bath immersion) were administered to twelve undergraduate participants in a randomized, crossover design. Five additional participants completed bath product treatment only. Participants collected saliva samples up to 48 hours after administration, which were analyzed for melatonin by enzyme-linked immunosorbent assay. Oral (*n*=11) and lotion formulations (*n*=12) caused maximum salivary melatonin levels within 30 minutes after administration, but bath immersion did not cause increases in saliva melatonin (n=17). The half-life of oral melatonin was 1.17 [0.69 – 1.65] hours versus 5.72 [3.75 – 7.68] hours for lotion treatment (*p* = 0.011, effect size *r* = 0.770). Melatonin lotion may pose a risk to consumers who assume it is safe and less effective than oral tablets, when in fact it may be very potent and remain at high physiological levels into the following day. This study is registered on clinicaltrials.gov (NCT06382610) and was funded by the Sleep Research Society.

**Statement of Significance:** We have shown that U.S. consumers assume that over-the-counter melatonin treatments like lotions and bath products are safe and less effective than oral tablets. Melatonin administered in bath product form does not result in melatonin absorbed through the skin. However, melatonin in lotion form contains a high level of the hormone which is absorbed readily and which lasts five times as long in the body than when taken orally. Lotions that are readily available in stores also have different ingredient lists, marketing claims, and amounts of melatonin which are not listed on the bottles. Consumers should be aware melatonin lotion may have a stronger impact than assumed.

## Introduction

The pineal gland’s natural production of melatonin promotes sleep and regulates circadian rhythm [1]. In the United States melatonin is not regulated as a drug by the Food and Drug Administration, whereas in countries like Canada, Japan, and the United Kingdom, melatonin must be prescribed by a physician [2]. The use of melatonin has been rapidly increasing in the last two decades in the United States [3]; sales in 2020 rose 42% compared to pre-pandemic 2019 sales [4] and increased another 53% between 2021 and 2024 [5]. This includes pediatric use. In a large U.S. cohort of 9 year olds followed for 6 years (*n* = 11,864) between 2016 – 2022, 12.2% of children used melatonin [6]; in a sample studied in 2023, 19% of 5 to 13 year olds (*n* = 543) had used melatonin in the previous 30 days [7] with parents only consulting a health care practitioner about half the time [8]. In the United States, over-the-counter transdermal melatonin products are available but often do not list dosages or instructions for quantity or frequency of use; this includes the “Dr. Teal’s” brand (PDC Brands, Stamford, CT), the top seller of mass market bath products [9]. A previous study showed that “Dr. Teal’s Sleep Lotion” contained 0.24% melatonin, or 2.4 ± 0.1 mg melatonin/g lotion, which increased salivary melatonin levels up to 1000-fold within an hour compared to a placebo lotion. Preliminary data also showed that the lotion improved sleep quality in participants with sleep difficulty [10]. However, half-life was not measured in that study nor was transdermal absorption compared to oral administration.

Knowledge of the pharmacokinetics of melatonin in different administration forms is important for effective use. For oral melatonin, the time to peak melatonin level in plasma (Tmax) has been reported as 15 -90 min and the half-life as 32 -126 min [11,12]. There have been limited studies of the pharmacokinetics of transdermal administration of melatonin, including patches [13,14] and melatonin mixed in 70% alcohol solution [15]. These small studies have not measured half-life but results point to a longer Tmax in plasma (1 – 13 h). The one study to our knowledge of melatonin in lotion form was a product formulated for the study with dimethyl sulfoxide (DMSO) as a carrier [16], which is not present in current commercial melatonin lotions. This administration (*n* = 10) resulted in a Tmax in plasma of 20.5 ± 8 h, a half-life of 14.6 ± 11.1 h, and a bioavailability of 10.0 ± 5.7%, but Zetner et al. did not compare this route to oral administration. The half-life of oral melatonin assessed via saliva has not been reported to our knowledge, and there has only been one study reporting Tmax in saliva as 60 min [17].

Without information about how well melatonin products are absorbed and how long they last in the body, consumers may be making mistaken assumptions about effectiveness as well as safety. Interviews with caregivers have identified a common belief that because melatonin is “natural” it will have fewer side effects [18]. Although melatonin is generally considered safe [19], new formulations would benefit from thorough assessments, particularly given its widespread usage in the United States.

In this study, we hypothesized that consumers would assume that transdermal melatonin products would be safer but less effective than oral melatonin tablets. We also hypothesized that melatonin in lotion and bath product form would show distinct pharmacokinetics compared to oral administration.

## Methods

### Consumer opinions

To examine U.S. consumer assumptions about the safety and effectiveness of melatonin products, we designed a survey approved by the University of Redlands Institutional Review Board (#2024-016) and in accordance with the Helsinki Declaration. We used G*Power 3.1 [20] to calculate that a sample size of 200 would yield 95% power to detect an effect size *f* of 0.13 with a within-participant correlation of 0.3. The survey was posted onto Surveyswap.com and Surveycircle.com in June and July of 2024 and participants provided informed consent. Participants (*n* = 199) were at least 18 years old and residents of the United States. Images of each of the following product bottles were shown to respondents: 3 mg melatonin tablet (Nature Made, West Hills, CA), Dr. Teal’s Body Lotion/Sleep Blend with Melatonin, Lavender, and Chamomile (PDC Brands, Stamford, CT), and Dr. Teal’s Foaming Bath/Sleep Bath with Melatonin, Lavender, and Chamomile (PDC Brands, Stamford, CT). Claims on the product label for the Nature Made tablet included “Supports a healthy sleep cycle and wake up refreshed” and “Gently ease into restful sleep.” Both the Dr. Teal’s lotion and bath products stated “Helps calm the mind for a good night’s sleep” on the labels shown. We asked survey respondents “How effective do you think each of these products is in improving sleep? We want your opinion on potential effectiveness, even if you have never used the product.” Participants could answer: not effective at all (1), a little effective (2), somewhat effective/unsure (3), moderately effective (4) or very effective (5). Similarly, they were asked “How safe do you think each of these products is? We want your opinion on potential safety, even if you have never used the product.” Possible answers were: very safe/little to no chance of harmful side effects (1), mostly safe (2), possibly safe/unsure (3), mostly unsafe (4), or very unsafe/high chance of harmful side effects (5).

Differences in effectiveness and safety ratings were assessed with Friedman tests followed by Wilcoxon Signed-Rank post-hoc tests due to non-normality of the data. The impacts of caregiver status (yes/no), gender (male/female), and ethnicity (Hispanic/non-Hispanic) were assessed with Mann-Whitney U tests. The impacts of age (collapsed into three categories for sufficient sample size: 18 – 24, 25 – 44, and ≥ 45 years) and race (collapsed into three categories for sufficient sample size: White, Asian, and other) were assessed with Kruskal-Wallis tests with post-hoc pairwise comparisons. All statistical analyses were performed using IBM SPSS statistical software (International Business Machines Corporation, Armonk, NY) with an alpha of 0.05.

### Melatonin quantification in lotion

Extractions of lotion samples in triplicate and High Performance Liquid Chromatography were performed as described previously, and our estimates of melatonin content in lotions assumed the same recovery from extraction (20%) as previously determined [10]. During the period of research for this publication, the Dr. Teal’s brand (PDC Brands, Stamford, CT) changed lotion formulations, label designs, and marketing claims. We tested three different types of products released between 2021 – 2025 that listed melatonin as an ingredient: 1) Dr. Teal’s Body Lotion/Sleep Lotion with Melatonin and Essential Oils in a 226.8 g tube, with the label claim “Promotes a better night’s sleep”, used in [10] (three lot numbers tested, including retesting the same tube used in that previous study); 2) Dr. Teal’s Body Lotion/Sleep Blend with Melatonin, Lavender, and Chamomile in a 226.8 g tube, with the label claim “Helps calm the mind for a good night’s sleep” (one lot number tested); and 3) Dr. Teal’s Body Lotion/Sleep Blend with Melatonin, Lavender, and Chamomile in a 532 ml pump bottle, with the label claim “Helps calm the mind for a good night’s sleep” (five lot numbers tested). All three formulations have differences in the ingredient lists on the labels. The first two product types no longer seem available. In a previous study [10] our laboratory tested Dr. Teal’s Body Lotion/Soothing Lavender Essential Oil, which has no label claims regarding sleep, as a presumed negative control and confirmed that it did not contain melatonin. This product no longer seems available. The current version of this product is called Dr. Teal’s Body Lotion/Soothe and Sleep Lavender Essential Oil with the label claim “Helps calm the mind to promote a better night’s sleep” but does not list melatonin as an ingredient. We tested this lotion as a presumed negative control.

### Melatonin quantification in bath product

We tested Dr. Teal’s Foaming Bath/Sleep Bath with Melatonin, Lavender, and Chamomile (label claim “Helps calm the mind for a good night’s sleep;” PDC Brands, Stamford, CT) as well as Dr. Teal’s Foaming Bath/Soothe and Sleep Lavender (label claim “Helps calm the mind promoting a better night’s sleep;” PDC Brands, Stamford, CT). The latter was a presumed negative control as the ingredient list does not contain melatonin. We were unable to analyze the bath products with the HPLC protocol used for lotions due to the concern of surfactants interfering with the column. Therefore, we attempted to quantitate melatonin in the bath products through fluorescence when excited at 280 nm (FP-8550 Spectrofluorometer, JASCO, Easton, MD). Standards were created in triplicate with 99% pure melatonin (ThermoFisher Scientific, Waltham, MA) in ultrapure water (>18 MΩ-cm obtained from a Barnstead E-PURE system, ThermoFisher Scientific, Waltham, MA) or methanol (VWR BDH Chemicals, Radnor, PA) from 0.1 ug/mL to 0.4 ug/mL. Bath products were diluted 1:10 to 1:200 for fluorescence analysis. Emission was analyzed from 290 – 450 nm, and peak emission from melatonin bath product occurred at 342 nm.

### Pharmacokinetics: randomized, crossover clinical trial

The study was approved by the University of Redlands Institutional Review Board (#2024-002) in accordance with the Helsinki Declaration and was registered on clinicaltrials.gov in April 2024 (NCT06382610). It is reported here following the CONSORT (Consolidated Standards of Reporting Trials) 2025 Statement with the previous extension to crossover trials [21,22]. The protocol and statistical analysis plan will be posted on clinicaltrials.gov upon publication. Deidentified participant data will be available in a public repository (TBD; will be provided on request) upon publication. A crossover repeated measures trial design was chosen to reduce the impact of between-subject variability; each participant underwent all three treatments and therefore serves as their own comparison. Treatments were separated by a 5 to 7 day washout period for each participant to prevent a carryover effect.

Participants were undergraduate students at a small liberal arts university in California, USA ages 18 – 21 years, not pregnant, not currently using melatonin, and without allergies or sensitivities to scented lotions or bath products. Recruitment occurred from May 2024 to July 2025. As calculated by G*Power 3.1[20] a sample size of 10 would yield 80% power to detect a 31 minute change in Tmax or half life with a within-participant correlation of 0.5, and a 25% change in peak melatonin level (Cmax) with a within-participant correlation of 0.25. We planned to recruit 12 participants to account for possible attrition. Twelve initial participants provided informed consent and were randomized by author LO via random number generator to sequentially numbered participants for the order of melatonin treatments (Figure S1). Treatments were not conducted blindly as it was impossible to conceal administration mode; research personnel were aware of the order of treatments after consent was obtained. Eleven participants received all three melatonin treatments; one participant dropped out of the study after a single treatment (lotion). Of the original 12 participants, 11 were female, 1 male; 8 were White, 3 “Other” race, and 1 more than one race; and 6 were Hispanic/Latino, 6 non-Hispanic/Latino. Participants who completed the study received a $50 USD gift card to a local retailer of general goods. Passive drool samples obtained with collection aids (Salimetrics, State College, PA) were frozen until analysis. Non-treatment saliva samples were taken at 2200h and 0400h to estimate the normal overnight rise in melatonin.

Pretreatment saliva samples were obtained at 1000h (0 hour timepoint) when endogenous levels of melatonin were anticipated to be low to provide a clearer comparison to exogenous treatment. Light levels were not regulated. Saliva samples and self-reports of side effects via questionnaire were obtained at 0.25, 0.5, 0.75, 1, 1.5, 2, 3, 4, 6, 8, 12, 18, and 24 hours after oral treatment. Transdermal treatments included these samples plus 30, 36, and 48 hour post-treatment timepoints, although not all samples were analyzed if melatonin levels had returned to baseline. All melatonin treatments were administered and the first hour of saliva samples were obtained under the supervision of research personnel. For oral administration, participants fasted for 2 h prior to and 2 h after swallowing a 3 mg Nature Made Melatonin tablet with 0.24 L water. For lotion administration, 3 g Dr. Teal’s Sleep Lotion with Melatonin, Lavender, and Chamomile (Lot number 23094 R C3603, PDC Brands, Stamford, CT) was applied to one arm and the area was not washed for at least 4 h. Given our HPLC analysis (Table 2), this amount of lotion is estimated to contain 3.9 mg melatonin. For bath product administration, participants soaked their feet and lower legs for 15 min in 35 L water maintained between 36.5 -40 C mixed with 1.5 ml Dr. Teal’s Sleep Bath with Melatonin, Lavender, and Chamomile (1:23,333 dilution); the exposed areas were not washed for at least 4 h. Five additional participants were recruited for a follow-up study using a higher concentration of sleep bath product; this extension of the study was not initially planned but was approved by the IRB prior to implementation. These participants included 3 females, 2 males; 4 White, 1 “Other” race; and 2 Hispanic/Latino, 3 non-Hispanic/Latino. Two participants were treated at 10- and 50-fold higher concentrations of bath product (1:2,333 dilution of product and a 1:467 dilution). The remaining three participants were treated with 50-fold higher concentration (1:467 dilution) only. These five participants received a $15 USD gift card to a local retailer of general goods.

Reagents for enzyme-linked immunosorbent assays to quantify melatonin in saliva samples were purchased from Salimetrics (State College, PA) and manufacturer instructions were followed. Optical density at 450 nm with reference at 620 nm was assessed using an Absorbance 96 plate reader (Byonoy, Hamburg, Germany). Standards from 0.78 to 50 pg/mL were used and extrapolation was allowed to 10% beyond the standard curve. Participant samples were diluted to 2x, 10x, 100x, or 1000x if necessary to fall within this range and all samples were tested in at least duplicate. Samples that fell under the lower limit of quantification (LLOQ) were assigned a concentration of the lowest standard (0.78 pg/mL) and one sample that was still not within the standard curve after thousand-fold dilution was conservatively assigned a concentration of 55,000 pg/mL. Out of 488 saliva samples tested, 4 were excluded as outliers. The average intraassay coefficient of variation was 3% and interassay coefficient of variation for this study was 10%.

Non-compartmental pharmacokinetic parameters were calculated using the program PKanalix (Lixoft-Simulation Plus, Research Triangle Park, NC). Timepoints analyzed for half-life were after Tmax and prior to the physiological increase of melatonin at nighttime (each curve was analyzed individually, and typically the last point included was 8 hours after treatment, at 1800h). Individual estimates for statistical analysis and time courses were generated using non-sparse data analysis. Missing data was omitted from analysis. Significant differences between oral and lotion treatment were analyzed on these individual estimates with sufficient data (*n*s = 10 – 12) using related samples Wilcoxon Signed Rank tests. Effect size *r* was calculated according to [23]. Overall administration mode estimates for Cmax, Tmax, and half-life were generated using the full data set with sparse data analysis. Exploratory analysis of relationships between pharmacokinetic parameters and body mass index were conducted with Spearman’s rho correlations. All statistical analyses were prespecified and performed using IBM SPSS statistical software (International Business Machines Corporation, Armonk, NY) with an alpha of 0.05.

## Results

To understand consumer assumptions about melatonin products, we surveyed adults in the U.S. (*n* = 199; Table 1) about their assumptions of effectiveness and safety of melatonin in oral tablet, lotion, or bath product formulations with rating scales from not effective at all (1) to very effective (5), and very safe (1) to very unsafe (5). Respondents assumed that oral tablets are more effective (score of 3.45 ± 1.05) than the transdermal products (lotion 2.36 ± 0.97, bath 2.36 ± 0.96; χ^2^(2) = 172.5, *p* < 0.001, effect size Kendall’s *W* = 0.43). Demographic variables were assessed for influence on ratings, and Asian respondents believed melatonin products in general were more effective (3.0 ± 0.73) than White respondents (2.68 ± 0.77, *r* = 0.29) or respondents of other races (2.43 ± 0.81, *r* = 0.23; χ^2^(2) = 9.12, *p* = 0.010). Gender, ethnicity, caregiver status and age had no relationship to effectiveness ratings (*p*s > 0.05, effect size *r*s <0.03, η^2^ =0.005). Respondents believed that all three forms of melatonin were equivalently “mostly safe” (oral 2.26 ± 0.99, lotion 2.25 ± 0.99, bath 2.22 ± 0.95; χ^2^(2) = 4.06, *p* > 0.05, Kendall’s *W* = 0.01), with no impact of gender, ethnicity, or caregiver status (*p*s > 0.05, *r*s < 0.09). White respondents assumed greater safety (2.10 ± 0.80) than Asian individuals (2.51 ± 0.94, *r* = 0.20; χ^2^(2) = 8.00, *p* = 0.018) and there was a trend for White respondents to assume greater safety than respondents of other races (2.41 ± 0.86; *p* = 0.07, *r* = 0.15). Asian respondents were not different than respondents of other races (*p* > 0.05, *r* = 0.04). Individuals ages 45 years and older assumed melatonin products were safer (1.94 ± 0.86) than did those ages 25 to 44 years (2.43 ± 0.80, *r* = 0.29) or 18 to 24 years (2.27 ± 0.85, *r* = 0.19); χ^2^(2) = 10.53, *p* = 0.005).

**Table 1.**
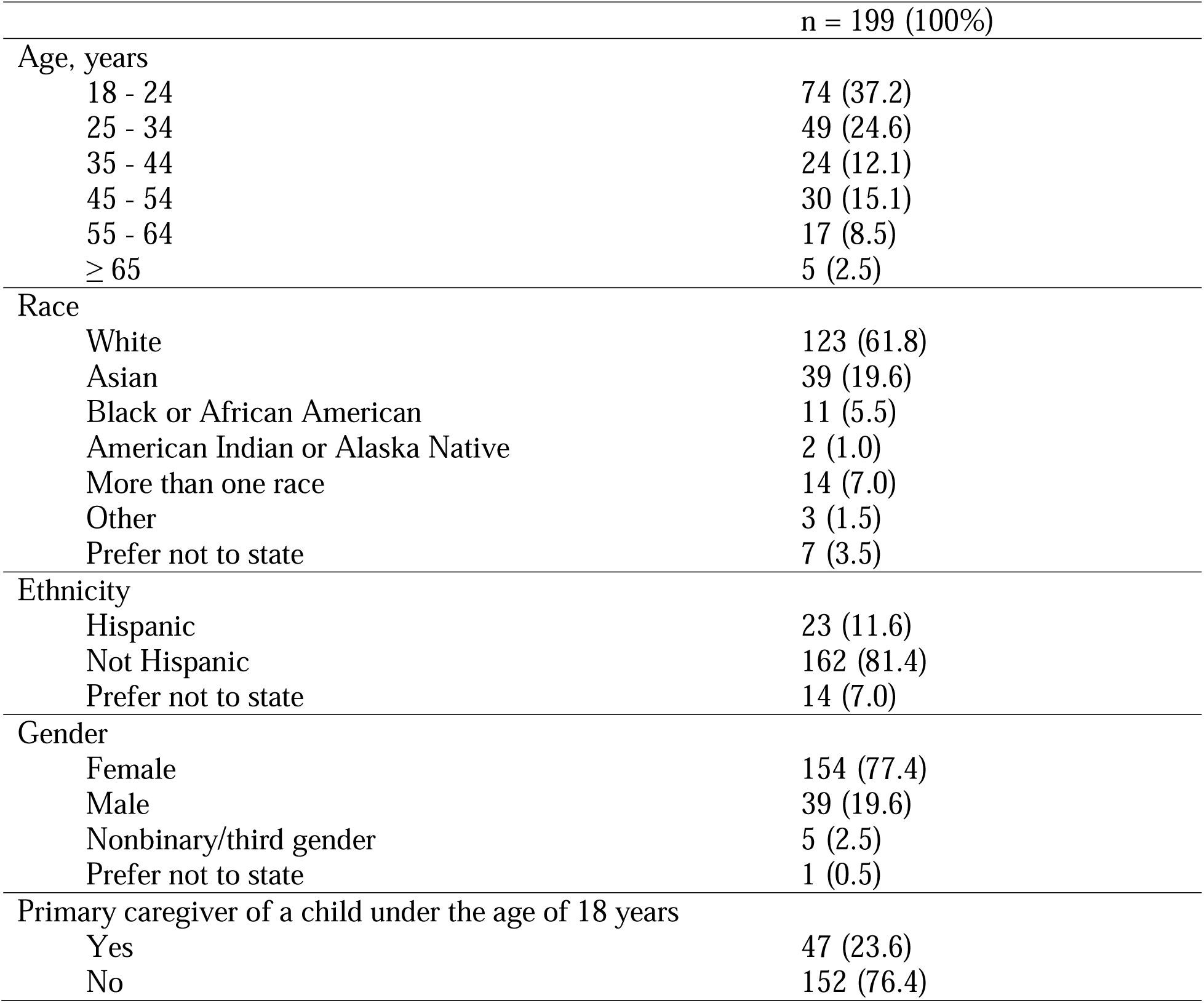
Demographics of respondents of consumer survey.

To confirm the presence of melatonin in Dr. Teal’s Body Lotion/Sleep Lotion with Melatonin and Essential Oils (PDC Brands, Stamford, CT) prior to the pharmacokinetics trial and assess lot-to-lot variability compared to a previous study [10], we performed High Performance Liquid Chromatography (HPLC) on lotion samples. It was discovered that three similar Dr. Teal’s brand melatonin lotion products were available over-the-counter with distinct ingredient lists on the labels and different marketing claims (Table 2). The consistency of the lotions differed qualitatively, with the two product types sold in 226.8 g tubes being thicker than the more liquid product sold in the 532 ml bottle. The lot-to-lot variability between products sold in tubes was also higher compared to the products sold in bottles: the standard deviation between newly assessed lots in tubes was 0.24 mg melatonin/g lotion (coefficient of variation of 17%) and the standard deviation between lots in bottles was 0.08 mg melatonin/g lotion (coefficient of variation of 8%). To assess degradation over time, one container of product that was assessed in July 2023 [10] was re-tested in July 2025. Potency was reduced by 54%.

**Table 2.**
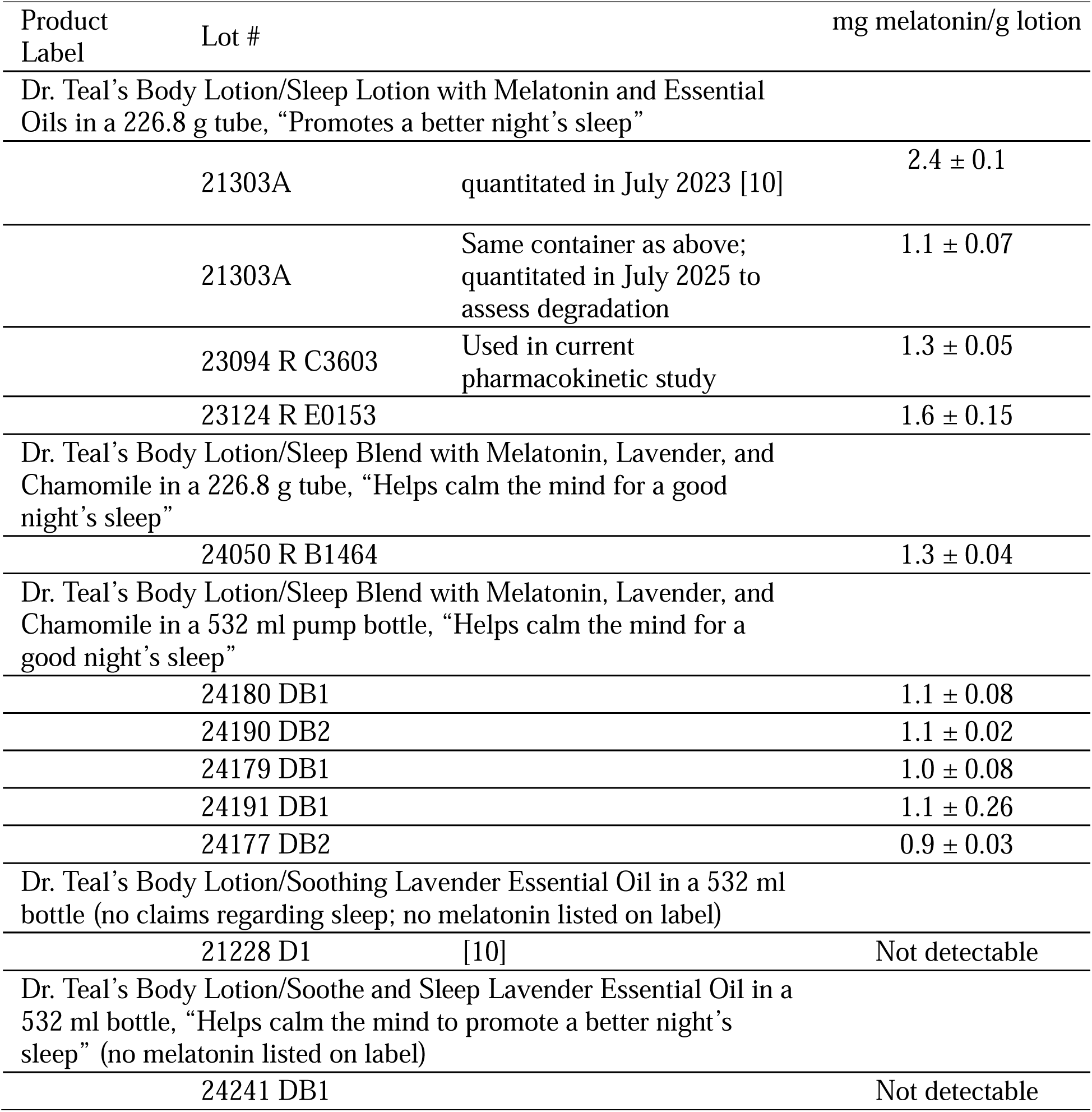
HPLC quantification of melatonin in Dr. Teal’s products.

Bath products could not be analyzed with the HPLC protocol used for lotions due to the concern of surfactants interfering with the column. Therefore, to confirm the presence of melatonin in Dr. Teal’s Foaming Bath/Sleep Bath with Melatonin, Lavender, and Chamomile (PDC Brands, Stamford, CT) we attempted to conduct fluorescence analysis via excitement at 280 nm and emission from 290 – 450 nm; the peak emission wavelength was 342 nm. We tested Dr. Teal’s Foaming Bath/Soothe and Sleep Lavender (PDC Brands, Stamford, CT) as a presumed negative control because the ingredient list does not contain melatonin, and we confirmed a lack of fluorescence at 342 nm. Although melatonin standards showed the expected linear relationship between concentration and fluorescence at 342 nm, samples of melatonin-containing bath product did not. Lower concentrations of bath product produced higher levels of fluorescence in both water and methanol diluents, suggesting a quenching effect of some component of the bath product. Thus we were unable to determine the quantity of melatonin in the bath product.

We conducted a randomized crossover clinical trial to determine the pharmacokinetics of melatonin absorption in oral tablet, lotion, and bath product form. We also assessed non-treatment melatonin levels at the timepoints most likely to include the physiological circadian rise (2200h and 0400h). Only melatonin in oral and lotion forms caused salivary melatonin levels to rise above pre-treatment levels (Fig 1). The only rise in salivary melatonin after bath treatment was equivalent to the physiological circadian rise. To determine if the lack of effect of melatonin bath treatment was due to insufficient concentration of bath product, we tested additional participants with 10-fold (*n* = 2) and 50-fold (*n* = 5) higher concentrations in a follow-up study. Salivary melatonin levels still did not rise above pretreatment levels, and all bath concentrations levels are combined in Figure 1.

**Figure 1.**
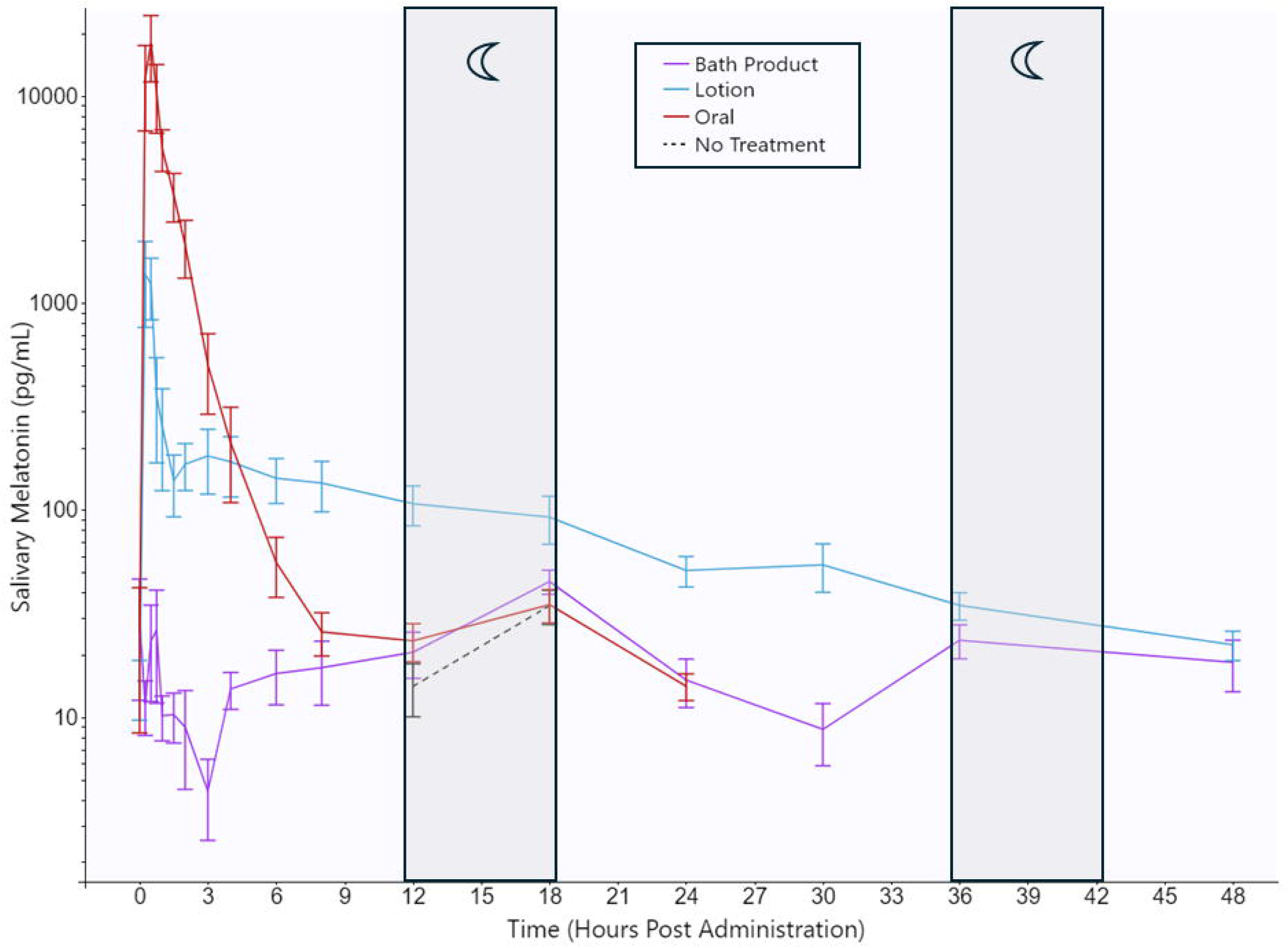
Salivary melatonin levels after oral, lotion and bath product melatonin treatments. Gray bars with moons indicate the 2200h to 0400h period (12 – 18 hours post administration) when endogenous melatonin was expected to rise. Only oral and lotion melatonin treatment resulted in salivary melatonin above physiological levels. Salivary melatonin after lotion treatment remained elevated for at least 36 hours. Note the y-axis log scale. Bath product treatment at all dosages (combined here) mimicked endogenous melatonin behavior, proving it to be ineffective even at 50x the original concentration. Error bars represent standard error. Alt text: Line graph with error bars with the x-axis from 0 to 48 hours post administration and the y-axis on a log scale up to 10,000 pg/ml salivary melatonin.

Since bath treatment did not influence salivary melatonin levels, we only calculated pharmacokinetic parameters for the oral and lotion treatments. Cmax was much higher after oral treatment than lotion treatment, although Tmax was the same (Table 3). The half-life of melatonin after lotion administration was approximately five times longer than after oral administration, and salivary melatonin levels were still above pretreatment levels 36 and 48 hours after lotion treatment. Exploratory analysis of relationships between pharmacokinetic variables and body mass index are shown in Table S1 although the sample size prohibits conclusions.

**Table 3.** Pharmacokinetic parameters after 3 mg oral melatonin and 3.9 mg transdermal melatonin lotion application.

|  | Oral Tablet<br>n = 11 | Lotion<br>n = 12 | p-value | Effect Size<br>(r) |
| --- | --- | --- | --- | --- |
| C<sub>max</sub> (pg/mL) |  |  |  |  |
| Sparse data analysis | 18,062 | 1,372 |  |  |
| Non-sparse data analysis |  |  |  |  |
| Mean [95% CI] | 20,791 [5,963 – 35,618] | 1,894 [653 – 3,135] | 0.004 | 0.86 |
| T<sub>max</sub> (h) |  |  |  |  |
| Sparse data analysis | 0.5 | 0.25 |  |  |
| Non-sparse data analysis |  |  |  |  |
| Mean [95% CI] | 0.61 [0.37 - 0.86] | 1.69 [0.20 – 3.18] | 0.238 | 0.36 |
| Half-life (h) |  |  |  |  |
| Sparse data analysis | 0.83 | 4.26 |  |  |
| Non-sparse data analysis |  |  |  |  |
| Mean [95% CI] | 1.17 [0.69 – 1.65] | 5.72 [3.75 – 7.68] | 0.011 | 0.77 |

The patterns of side effect symptoms did not appear to differ between oral, lotion, and bath melatonin treatments (Table 4), although the small sample size prevents statistical analysis and there is no placebo comparison. No participants reported skin irritation due to transdermal treatments.

**Table 4.** Number of individuals reporting side effects after melatonin treatment.

| Melatonin<br>administration form | Drowsiness | Headache | Vivid<br>Dream/<br>Nightmare | Dizziness |
| --- | --- | --- | --- | --- |
| Oral tablet | 5 | 3 | 1 | 1 |
| Lotion | 5 | 2 | 4 | 1 |
| Bath product | 4 | 3 | 2 | 1 |

## Discussion

We found that U.S. consumers assumed that oral melatonin products are more effective than transdermal melatonin products but that they are no different in safety. In analysis of pharmacokinetics, melatonin administered in lotion form had a significantly longer half-life than oral administration, but bath product administration had no effect on salivary melatonin levels at all.

Lotion products, which are unregulated as drugs in the U.S., somewhat differed in melatonin content between products with very similar packaging. Unlike analyses of other melatonin products such as tablets and gummies [24–26], we cannot compare our estimates of melatonin content to claimed dosages since there were none listed on packaging. However, with melatonin content ranging from 0.9 -1.6 mg/g lotion in this study, it is likely that consumers are applying substantial doses (>10 mg).

Oral melatonin resulted in a significantly higher (∼12-fold) Cmax than melatonin lotion in the present study, with a similar Tmax. The dosage of the oral tablet was 3 mg as listed on the label. In a previous study, the application of 7 g Dr. Teal’s Sleep Lotion (lot 21303 A, a dose of 16.8 mg melatonin) resulted in a Cmax of 16,392 [95% CI 10,602 – 22,183] pg/ml salivary melatonin 30 min after treatment [10]. Since saliva samples were only obtained at 30 min and 1 hour after treatment, half-life was not determined in that study. Given the high levels of melatonin absorbed and lack of knowledge of half-life, we chose to administer 3 g Dr. Teal’s Sleep Lotion in the present study (lot 23094 R C3603, a dose of 3.9 mg melatonin). This resulted in an approximately 10-fold lower Cmax than in [10], although it would still be 10 to 50-fold higher than expected overnight physiological levels. A typical consumer would probably apply more than 3 g lotion and would therefore be receiving a potent dose.

Melatonin administered in bath product form did not alter salivary melatonin levels in our participants. Our mock bath protocol only immersed the feet and lower legs of participants for 15 minutes, which is less exposure than expected in an at-home bath. However, to test the hypothesis that participants had been insufficiently exposed to the product, we conducted follow-up studies using 10-fold and 50-fold more concentrated solutions. At the 50-fold higher concentration, this would scale up to adding ∼320 mL product in a standard-size bathtub holding 150 L, a concentration substantially higher than we expect consumers would use. Instructions on the bottle say only “pour a generous amount” into the bath. Although there is still the possibility that exposure to a larger surface area would cause salivary melatonin levels to rise, we hypothesize that the formulation used in the bath product or the dosage of melatonin is not compatible with transdermal absorption.

We were unable to quantitate the amount of melatonin in the bath product by either HPLC (due to surfactants) or fluorescence analysis (due to a quenching effect, where increased sample concentration yielded lower fluorescence intensity). It is unknown what components in the bath product might, at higher concentrations, reduce the intensity of melatonin fluorescence. Melatonin fluorescence is known to be quenched by aerated oxygen and by halogenated molecules [27]. Fluorescence intensity of melatonin is also decreased in natural deep eutectic solvents, with components such as fructose, glucose, citric acid, lactic acid, glycerol, and menthol [28]. In contrast, melatonin fluorescence intensity increases with increasing hydroxypropyl β-cyclodextrin concentration as well as with the micelle-forming surfactants sodium dodecyl sulfate and cetyl trimethyl ammonium chloride [27]. Further research such as the isolation of melatonin from other components of the bath product may allow for its analysis via fluorescence.

Side effects reported in this study would be considered nonserious adverse events and were similar to those previously described [19,29,30] including drowsiness, headache, vivid dream/nightmare, and dizziness. The small sample size and lack of placebo control preclude statistical analysis, but it is interesting to note that similar side effects were reported with bath product exposure which had no impact on salivary melatonin levels.

Limitations of our pharmacokinetics study include a gender skew in recruitment, a restricted age range, and a small sample size that limits full analysis by body mass index, demographic variables, and of side effects. Although carryover effects are a potential issue with a crossover trial design, our washout period of 5-7 days between treatments should have been sufficient to prevent this given our half-life results. A limitation in the HPLC studies is that we are assuming the same recovery of melatonin (20%) in the extraction process in different formulations and over time as previously determined [10], thus our quantification should be considered an estimate. For our survey of consumer opinions, our sample consisted of participants on Surveyswap.com and Surveycircle.com, which are websites that facilitate the exchange of participation in research studies. Thus, respondents are typically designers of their own surveys and are probably more educated than the general population.

Understanding pharmacokinetics of new administration forms is necessary for informed use of melatonin. This is particularly urgent given increasing melatonin usage in the United States even in pediatric populations; for example, 2023 survey data showed that 1.7% of parents had administered melatonin (mostly in gummy form) to infants/toddlers under the age of 1.5 years old [31]. Transdermal products are probably appealing to parents of young children especially. We have shown that consumers assume that transdermal products are safe and less effective than oral tablets. In light of this, the potency and prolonged half-life of melatonin administered in lotion form is worthy of attention.

## Data Availability

All data produced in the present study are available upon reasonable request to the authors.

## Acknowledgements

The authors would like to thank Dr. Michael Ferracane, Dr. Teresa Longin and Dr. David Schrum for consultation on fluorescence analysis and HPLC.

## Disclosure Statement

Financial Disclosure: none.

Non-financial Disclosure: none.

A preprint of this manuscript is available from medrxiv.org.

## Supplemental figure legend

Figure S1. CONSORT (Consolidated Standards of Reporting Trials) participant flow diagram for pharmacokinetics clinical trial (clinicaltrials.gov NCT06382610).

Alt text: Flow chart showing the number of participants at each stage in the clinical trial.

