## Supplementary figures and images for "Consumer Opinions, Lot-to-Lot Variability, and Pharmacokinetics of Transdermal Melatonin Products: A Randomized, Crossover Clinical Trial"

### Fig S1

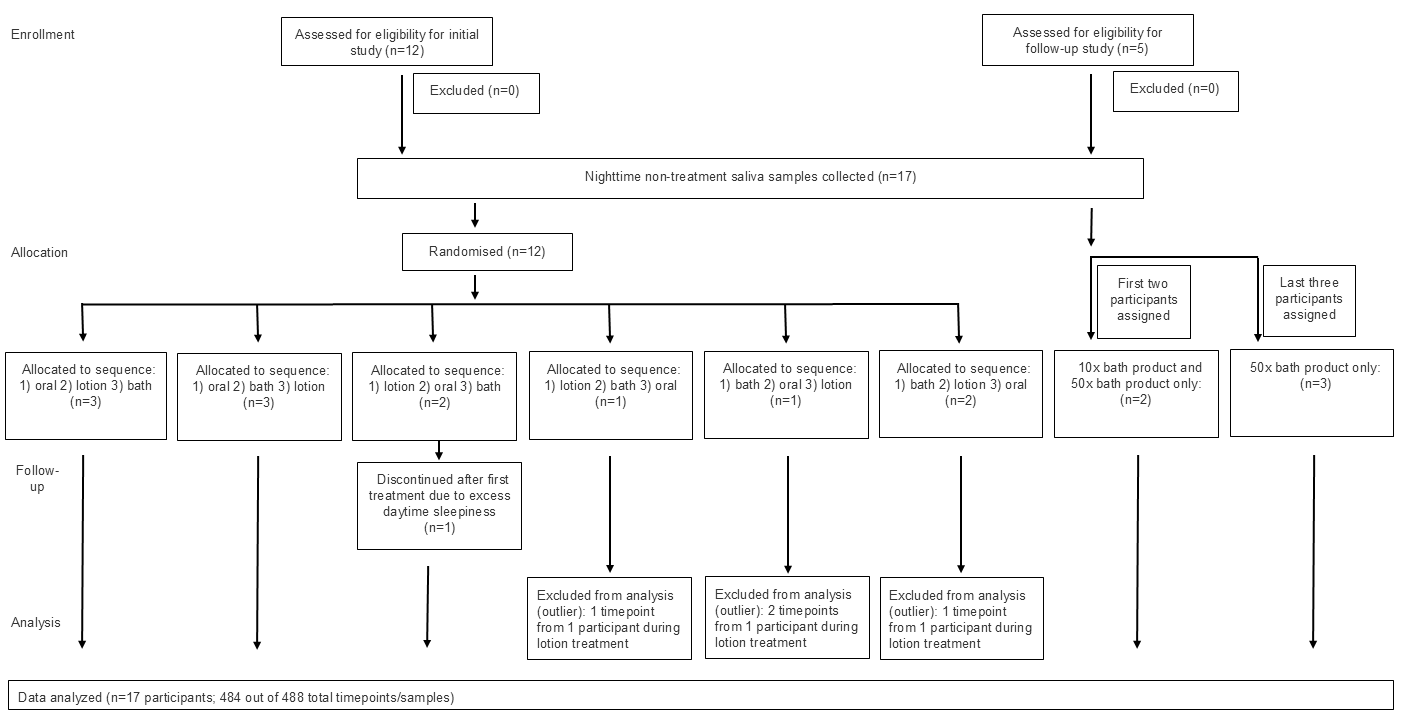
