## Supplementary material for "Consumer Opinions, Lot-to-Lot Variability, and Pharmacokinetics of Transdermal Melatonin Products: A Randomized, Crossover Clinical Trial": Table S1

Table S1. Exploratory analysis of pharmacokinetic parameters and body mass index

|  | Cmax Oral | Cmax Lotion | Tmax Oral | Tmax Lotion | Half-life Oral | Half-life Lotion |
| --- | --- | --- | --- | --- | --- | --- |
| Body Mass Index  ρ (Spearman’s rank correlation coefficient) | -0.04 | 0.18 | -0.05 | -0.17 | -0.29 | 0.38 |
| *p* | 0.92 | 0.57 | 0.89 | 0.60 | 0.39 | 0.28 |
| *n* | 11 | 12 | 11 | 12 | 11 | 10 |
